# Brazilian dental students and COVID-19: a survey on knowledge and perceptions

**DOI:** 10.1101/2020.07.28.20163964

**Authors:** Maria Gerusa Brito Aragao, Francisco Isaac Fernandes Gomes, Letícia Pinho Maia Paixão de Melo, Silmara Aparecida Milori Corona

## Abstract

This study evaluated the knowledge and perception of Brazilian dental students about COVID-19 and the undergraduate clinical practice during the outbreak by a self-administered web-based questionnaire. A social network campaign on Instagram was raised to approach the reach population. The survey covered demographic and academic profile, general knowledge, preventive measures, and perception about the COVID-19. Descriptive statistics were used to identify frequencies and distributions of variables, which were compared by type of institution and current year of enrollment using Chi-square or Fisher’s exact tests (α=0.05). A total of 833 valid responses were received over 10 days. Students were able to identify the incubation period, main symptoms, and contagious routes of the disease but struggled in recognizing the name of the virus responsible for the pandemics. Hand washing before and after a dental appointment with a patient (97.7%) followed by use of barriers to protect mucosa (97.2%) were the more frequently recognized measures to prevent COVID-19 spread in the dental office. As for the perception of COVID-19, 73.2% of the dental students perceive the disease as severe, while only 11.1% of them think that COVID-19 is severe only for people presenting risk factors. Dental student’s knowledge and perception were associated with the type of institution and year of enrollment. In summary, the dental students demonstrated an acceptable general knowledge about COVID-19, but dental schools will need to address gaps in knowledge and control measures and perceptions to ensure a safer return to presential activities.

## Introduction

COVID-19 is a novel disease caused by a member of the coronavirus family SARS-Cov-2 that originated in Wuhan, Hubei, China in December 2019 (Gralinski & Menachery, 2020; Wang et al., 2020). Over a few months, it became a pandemic that has globally affected over 13 million people, causing more than 690,000 death episodes in 216 countries according to the World Health Organization (WHO) (WHO Coronavirus Disease Dashboard,2020). Brazil became a COVID-19 epicenter in Latin America with rocketing numbers of cases of COVID-19, as well as death episodes over this period. By July 2020, Brazil ranked number 2 globally in the prevalence of COVID-19, reaching approximately 2.8 million cases, whereas death episodes rose above 97,000 cases (WHO Coronavirus Disease Dashboard, 2020).

The pandemics imposed a heavy toll to basic human activities and social distancing is by far the most effective measure to ease off further spread of COVID-19 among individuals once vaccination and therapeutic approaches are still under development (Raimondi et al.,, 2020; Omolo et al., 2020; Qian & Jiang, 2020). Amid this chaotic scenario, pandemics are also a huge challenge to healthcare professionals who are at high risk of being contaminated due to frequent exposure to contaminated patients (Adams & Walls, 2020; Li et al., 2020). Among these healthcare workers, dental professionals are at the top of the pyramid of professionals at risk, since the dental practice involves face-to-face communication with patients and frequent exposure to oral fluids and the respiratory system (Prati et al., 2020; Ghai, 2020). The clinical environment thus became critical given the aerosols produced by rotatory equipment, as well as due to oral fluid and tissue manipulation 9Meng et al., 2020; Peng et al.,2020).

In Brazil, the universal dental care covers approximately 211 million people distributed along an 8.5-million-km^2^ area and approximately 330,000 dentists are in the workforce (Moraes et al., 2020). Most of these professionals had their work status affected, especially in less developed regions. The impact on routine was high for them, leading to varied changes to clinic infrastructure, personal protective equipment use, patient screening, and increased costs (Moraes et al., 2020). During the COVID-19 outbreak, as these professionals are in such a vulnerable position, dental students also might be facing several challenges to cope with the current pandemics (Prati et al., 2020; Meng et al., 2020; Desai, 2020; Iyer et al., 2020; Liu et al., 2020; Machado et al., 2020; Wu et al., 2020). Brazil counts on many tertiary institutions responsible for formal dental education at a graduate level. According to the last census, 350 institutions offer dentistry as a formal degree to over 125,585 students across the country^19^ (Brazilian National Institute of Educational Research and Studies [INEP], 2019). In the context of COVID-19, social distancing, the university lockdowns, and the high risk of contamination during clinical practice, dental education finds a stumbling block ahead to deliver the hands-on experience to dentistry students in the near future, assuming there continues to be an infection risk in the foreseeable future(Meng et al., 2020; Iyer et al., 2020; Liu et al., 2020; Machado et al., 2020).

Many institutions have issued protocols to avoid further spread of COVID-19 (WHO Clinical management of COVID-19 Interim Guidance, 2020; Brazilian Federal Dental Board, 2020), many experts in the field of dental education have discussed the future of dental education from an institutional perspective (Desai et al., 2020; Liu et al., 2020; Machado et al., 2020; Wu et al., 2020), and some studies have dwelled on the professional’s awareness and attitudes towards COVID-19, which are essential reports on this serious issue. Yet, at least in Brazil, not so many studies have questioned dental students their awareness towards this pandemic scenario, as well as how they perceive the disease concerning dental education, which has been done elsewhere (Van Doren et al., 2020). Thus, this study aimed to evaluate the knowledge and perception of Brazilian dental students about COVID-19 and the clinical practice in the context of the pandemic.

## Methods

### Ethical aspects

This research protocol was approved by our research ethics committee (CAAE: 33608320.5.0000.5419). The study consists of a cross-sectional survey directed to a sample of dental students.

### Questionnaire preparation

A self-administered questionnaire about the awareness and knowledge of dental students about COVID-19 and its impact on the undergraduate dental practice was developed based on the Centers for Disease and Control (CDC) Guidance for Dental Settings (CDC Guidance for Dental settings, 2020) and the Manual of Good Practices and Biosafety of the Brazilian Federal Board of Dentistry (Brazilian Federal Dental Board, 2020). The Clinical Management of COVI-19 Interim Guidance of the WHO (WHO Clinical management of COVID-19 Interim Guidance) was also consulted. The questionnaire was hosted online (Google Forms). A pre-test was conducted with 10 undergraduate dental students to evaluate the validity of the research tool chosen and of the questions. In the pre-test, the students were asked to evaluate the questions regarding their clarity. The first and last questions of the pre-test were “What time is it?”, so we could calculate the meantime the students took to answer the questionnaire. In the pre-test, the students scored the clarity of each question on a scale of 1 (not clear) to 5 (very clear). As all items scored ≥ 4, so we did not change the way the questionnaire was organized. The mean time to complete the survey ± standard deviation (SD) was 9±3 min. Those who participated in the pre-test were excluded from participating in the main study to avoid response bias.

### Questionnaire design and content

The participant had to click ‘Yes’ after the question “Do you agree to participate in the study ?”. The questionnaire contained 20 mandatory close-ended items, divided into four sections: demographic and academic profile (n=6); general knowledge about the COVID-19 (n=4); knowledge about the preventive measures to avoid COVID-19 spread on the undergraduate dental practice (n=2); and perception about the COVID-19 impacts on the undergraduate dental courses (8). The options ‘I'd rather not say’, ‘I don't know how to answer’ were treated as missing data. This manuscript does not cover the entire questionnaire.

### Online survey

We used the data of the last Brazilian Tertiary Education Census (INEP - Brazilian National Institute of Educational Research and Studies, 2019) to know the size of our source population. According to this census, there are 125,585 undergraduate students enrolled in dentistry courses in Brazil considering public and private institutions. All these students were eligible to participate in the research. Considering the target population of 125,585 dental students, we estimated that 693 responses would be necessary to ensure a 95% confidence interval and a 3% margin of error, also considering 20% of possible losses.

To recruit the participants, we created an Instagram® social networking campaign targeting dental students in Brazil (Facebook, Menlo Park, CA). We followed the strategy described by Moraes et al., 2020^13^, who performed a national survey directed to Brazilian dentists. Thus, an Instagram professional account was created (@covid.forp) with a website link to the questionnaire in its bio page. First, we created one post with a text image containing only the title of the research. In the post description, we provided the main objectives of the project, called for participants, and asked them to share our post. Dental students with professional Instagram accounts were asked to share our post on their stories to increase the reach to the target population and to assist in disseminating the campaign. Five days after the first post was created, we made a second post with a different visual aspect. In this second post, we included in the image the title of the research project and a phrase asking people to share our post in their stories. Brazilian dental students, professors, and Instagram profiles of dental schools shared our posts. The campaign started on July 4 and lasted until July 14.

### Data analysis

The data collected was extracted from Google Forms and converted to Excel (Microsoft, USA) sheets. Descriptive statistics were used to identify frequencies and distributions of variables. The frequencies distribution for the general knowledge about COVID-19, knowledge about the preventive measures to avoid COVID-19 spread on the undergraduate dental practice, and perception about COVID-19 were compared by type of institution (public versus private) and by current year of enrollment (first to fifth year) using Chi-square or Fisher’s exact tests and a significance level set at 5%. All statistical tests were performed using the 7.0 GraphPad Prism® software (California, USA).

## Results

A total of 833 valid responses were received over 10 days (Fig. 1) from participants from all Brazilian states and the Federal District. Participants were mostly female (80.1%), aging between 18 and 25 years old (85.1%), and studying dentistry at institutions located in the Southeast of Brazil (54.7%). Moreover, 51% of the respondents studied dentistry at private institutions and most of them were enrolled in the fourth (24.8%) and fifth (25.3%) years of the undergraduate course (Table 1).

When asked the name of the virus responsible for the pandemics we are facing, 50.1% of the participants answered correctly, choosing the option SARS-Cov2, whereas 40.8% chose the option COVID-19 (Table 2). When analyzing these data by type of institution, it was observed a significant difference in the distribution of frequencies. While 64.9% of students from public institutions chose the option SARS-COV2, 52% of students from private institutions chose COVID-19 (Table 3). The same data analyzed by year of enrollment showed that second-year students identified the right name of the virus more frequently (52.9%), yielding a statistically significant difference. At the same time, third-year students were the ones who chose the option COVID-19 more frequently (44.6%) (Table 4).

Regarding the incubation period, 67.2% of respondents considered 7 to 14 days to be the SARS-Cov2 incubation period (Table 1). No statistical difference was observed when comparing the response frequencies of students from public and private institutions (Table 3) or different year of enrollment (Table 4). Contact with air droplets was considered by the dental students the main contagious route of COVID-19 (95.2%), while only 1.7% chose the option accidents with sharp objects as a possible contagious source (Table 1). There was no statistical difference when analyzing these data by type of institution (Table 3) or by year of enrollment (Table 4). The dental students recognized difficulty breathing (95.6%) and fever (95%) as the main symptoms of COVID-19 (Table 2) No association with the type of institution was found (Table 3) but significant higher proportions of students from the last years of the dentistry undergraduate course recognized dry cough as the main symptom of COVID-19 (Table 4).

Hand washing before and after a dental appointment with a patient (97.7%) followed by the use of barriers to protect mucosa (97.2%) were the more frequently recognized measures to prevent COVID-19 spread in the dental office. The use of mouthwashes (30.5%) and the use of rubber dam (32%) were the less frequently chosen preventive measures (Table 2). Comparing data of students from public and private students, a significant difference was found among proportions. In this case, the differences were observed in the less frequently chosen options, 38.6% of students from public dental schools recognized the use of manual instruments as measures to prevent COVID-19 spread, an option that was chosen only by 24% of students from private institutions (Table 3). No statistical difference among proportions was found when these data were analyzed by year of enrollment (Table 4).

In terms of biosafety training to deal with dental practice during the COVID-19 outbreak, only 4.1% have received practical training, 42.3% have never been trained, and 53.5% of students have received only general information without practice (Table 2). Higher proportions of students from private institutions have received general information without practice (56.5%), while the same was true only for 50.0% of dental students from public universities, yielding a significant difference. Moreover, while 6.9% of students from private institutions have received practical training, the same was observed only for 1.3% of students from public universities (Table 3). No statistical difference was found when these data were analyzed by year of enrollment. (Table 4)

As for the perception of COVID-19, 73.2% of the dental students perceive the disease as severe, while only 11.1% of them think that COVID-19 is severe only for people presenting risk factors (Table 2). Such perception was associated with the type of institution since higher proportions of students from public institutions perceived COVID-19 as a severe condition (Table 3). This perception was not associated with the year of enrollment (Table 4). The students (86.2%) also considered that there is a high risk of infection and transmission of COVID-19 in the clinical practice at the dental schools (Table 2). Higher proportions of students from public dental schools perceived the risk of infection as high (94.7%) (Table 3) but no association with the year of enrollment on such perception was found (Table 4).

Concerning the impact of the pandemics on the dental school routine, 71.1% of students considered it to be very strong and only 0.2% considered that the outbreak of COVID-19 caused a low impact on the dentistry undergraduate courses (Table 2). Significative higher proportions of students from public dental schools (75.8%) considered the impact of the pandemics as very strong (Table 3). Such perception was also associated with students from the fourth (79%) and fifth year (73%) of dental school (Table 4). When asked about their feelings regarding returning to dental school, 32.6% of the students responded that they were completely worried, while only 6.9% answered that they were not worried (Table 2). This feeling was associated with the type of institution as a higher proportion of students from public dental schools chose the option “I am completely worried, while students from private dental schools answered that they were only worried (Table 3). The concern regarding returning to the dental school routine was not associated with the year of enrollment. Even being worried about returning to the dental school activities, 92.7% of students would not change their degrees, an opinion that was more frequent in students from private dental schools (96%) than in the public ones (89.1%) (Table 3), regardless of the year of enrollment (Table 4).

Finally, most of the students (78%) have never been suspected or diagnosed with COVID-19, and only 3.3% of them have tested positive for the disease (Table 2), a result that was associated with students from public dental schools, who have never been suspected or diagnosed with the disease (82.3%) (Table 3) but not with the year of enrollment (Table 4). Higher proportions of students (77.3%) responded that they do not have risk factors for COVID-19, while 6.3% of them responded they do not even know it (Table 2). COVID-19 suspect was significantly more common on students from public dental schools (82.3%) than from private (74.6%) (Table 3), regardless of the year of enrollment (Table 4).

## Discussion

COVID-19 has challenged schools worldwide, and the higher education sector has not been immune to it (Sahu, 2020, United Nations Educational, Scientific and Cultural Organization [UNESCO], 2020). Healthcare professionals and students have been strongly affected by the need for social distancing and the consequent closing of their schools (Adams et al., 2020; Quinn et al., 2020). Dentists, as well as dental students, are at the top of the pyramid of healthcare professionals at risk (Meng et al., 2020). Thus, the continuity of activities in dental schools during the COVID-19 outbreak will face practical and logistical challenges and concerns for patient safety once students may potentially spread the virus when asymptomatic and may acquire the virus in the course of training (Ghai, 2020; Iyer et al., 2020; Quinn et al., 2020). In Brazil, returning to direct patient care will be a huge challenge for 350 institutions that offer dentistry degrees to over 125,585 (INEP, 2020) students once the country has become one of the epicenters of the disease. Presential activities will require extensive preparation, and the success in transitioning will be associated with the students’ preparedness to deal with the patient care routine (Prati et al., 2020; Ghai, 2020, Quinn et al., 2020). The students’ basic understanding of the disease will be a start point for dental schools to plan their return. Therefore, here, we investigated the knowledge, awareness, and perception of Brazilian dental students about clinical practice in the COVID-19 context.

To reach dental students, we made an Instagram campaign, following the methods described by Moraes et al., (2020). After the 10-day campaign, we received 833 valid responses, and, to the extent of our knowledge, this is the first study in Brazil to recruit such a large sample of dental students in a web-based survey about COVID-19 knowledge and perceptions. In Brazil, Instagram is a largely used social media. In July 2020, Brazil figured as number 4 country in the reach rankings as one of the countries with higher numbers of access and audience increase (We are social, 2020). Moreover, Instagram is highly used by dentists and dental students in Brazil, which might have facilitated our recruitment. For instance, on July 20 there were 6.9 million posts using #odontologia, 2.8 million using #odonto, and 40.6 thousand posts using #estudantedeodontologia (The first two are the Portuguese words for dentistry and the last one is the Portuguese word for dental students). Thus, in our study, Instagram was a resourceful tool for the survey spread and divulgation.

As for the use of an online survey, it is known that it presents limitations (Couper, 2001). However, given the conditions imposed by the pandemics, web-based surveys allow for data collection without disrespecting sanitary measures. Moreover, such surveys present low costs and broad access to subjects, allowing large-scale data collection and processing in a relatively small amount of time (Couper, 2001). In terms of response rates by gender, we observed a proportion comparable to the overall gender distribution of dental students in Brazil, where dentistry is among the ten undergraduate courses with higher proportions of women (INEP, 2019). In terms of participation by Brazilian regional division, we received more than half of our responses from students from dental schools from the Southeast of Brazil, which is where the research was originated and also is the region with higher proportions of dental students, accounting for 43.8% of the total (INEP, 2019). Lastly, dentistry undergraduate courses in Brazil are mostly offered by private institutions, which supports the slightly higher participation of students from private dental schools (INEP 2019)

Regarding the dental students’ general knowledge about COVID-19, our results show that while students were able to identify the incubation period, main symptoms and contagious routes of the disease, they struggled in recognizing the name of the virus responsible for the pandemics we are facing in 2020, which is a worrisome problem. As it is shown in Table 2, more than 40% of the participants confused the name of the virus with the name of the disease. This misunderstanding shed light on the necessity of schools to prepare their educational materials to provide the students with reliable information (Ghai, 2020; Desai, 2020). To aid in this task, Elsevier (Elsevier COVID-19 Health Education resource Center, 20200 and the Food and Drug Administration (FDA) (FDA covid-19 Educational Resources) have created an online platform with COVID-19 Health Education resource centers, where they have launched podcasts, videos, summaries, and skills designed to support the learning of healthcare students. Moreover, there is a range of educational materials available in Portuguese on the UNA-SUS website (UNA-SUS, 2020), which consists of a platform organized by the Brazilian Ministry of Health to provide continuing education for health students and professionals. Special attention must be given to students from private dental schools, who were the ones who confused the name of the virus with the name of the disease more frequently. As in Brazil there are around 100,000 (INEP, 2020) thousand students enrolled in private dental schools, they must acquire basic knowledge on the general aspects of COVID-19 before returning to the clinical activities. The same applies to students in the last years of the undergraduate course, who will soon be working with patient care on their own.

When looking at our results regarding the students’ knowledge about measures to prevent the risk of COVID-19 spread, we observed consistency between their choices with the recommendations of the already published guidelines on biosafety during the COVID-19 outbreak (Brazilian Federal Dental Board, 2020; CDC Guidance for Dental Settings, 2020). The current dental curriculum at most dental schools addresses basic infection control, mainly focusing on preventing blood-borne infections, such as HIV and hepatitis (Ghai, 2020). In Brazil, before the pandemics, the federal dentistry board periodically released biosafety guidelines that are adopted by dental schools over the country. However, airborne and droplet infections appeared to be seldom addressed. In the pandemic context, the federal board has released a new guideline available in Portuguese including precautions regarding diseases such as H_1_N_1_ and COVID-19 (Brazilian Federal Dental Board, 2020). Attention must be also directed to the changes that COVID-19 has brought to patient care and operative dentistry. Due to possible aerosol transmission of SARS-Cov2 (Ge et al., 2020) the use of high-speed dental piece must be avoided and manual instruments must be chosen instead (Meng et al., 2020; Ge at al., 2020). Preventive and minimally invasive dentistry should be preferred, and the use of rubber dam must be mandatory when high-speed pieces are used (Ge et al., 2020; Peng et al., 2020). Our results also showed that few respondents have received practical training regarding preventive measures, which emphasizes the need for workshops and hands-on activities before returning to activities at dental schools (Ghai, 2020).

Most of the participants did not show good perceptions regarding COVID-19, mainly the ones from public dental schools or the ones in the last years of the undergraduate course. Participants mostly perceived COVID-19 as a severe disease, classified the risk of infection as high, and considered the pandemics to have strongly impacted on dental schools. In this sense, we can discuss that teaching and learning have been profoundly affected (Prati et al., 202; Wu et al., 2020; Van Doren et al., 2020), most of the activities have shifted online, and the biggest challenge is to postpone direct patient care, as no virtual sessions can duplicate the clinical experience (Desai, 2020; Machado et al., 2020). We also stress that more investigation needs to be performed on how social distancing and remote learning has affected dental education and how to maintain high-quality teaching and learning if the scenario persists to be risky. As graduation ceremonies have been canceled or delayed, the participants are also very worried regarding returning to presential activities. In Brazil, this perception might be associated with the rise of cases and deaths, which reinforces the need for dental schools to consider the students’ general anxiety and safety concerns. In this scenario, it would be important to know if students present risk factors for COVID-19 or have ever been suspect or diagnosed with the disease (Ghai, 2020). In our study, most respondents have never been suspected neither diagnosed with the disease nor do they present risk factors. This information is valuable when planning the return to presential activities. Thus, we suggest that each dental school must approach their students individually to better understand the scenario the school might face and to ensure screening and low-risk exposure. Even with all the difficulties imposed by the pandemics on dental education, most of the respondents would not change their degree, which implies they are still motivated to continue their education.

As for the limitations of this study, we point that rejection and losses cannot be estimated. Other limitation regards the representativeness of the sample of responders, as the data analysis did not consider ethnicity and income of the sample. The results might not be generalized, yet they are representative of the target population. Given the current pandemic situation, responses might present bias. Moreover, the sample recruitment using only social media might have limited the reach of our research and maybe combined strategies, such as use of email and web-based surveys would have increased our spread though difficult to reach Brazilian regions. Reding strengths, our study considers a population of students highly affected by the COVID-19 pandemic. We addressed their knowledge and perceptions, which are important information for planning strategies in clinical dental education during the outbreak. Moreover, our questionnaire might be a tool to dental schools to assess the scenario they are going to face when students are back to practice and in which aspect they will have to focus.

## Conclusions

Although dental students demonstrate an acceptable general knowledge about COVID-19, some aspects of the disease and control measures need to be better addressed by dental schools to ensure a safer return to presential activities. Moreover, as dental students did not present good perceptions about COVID-19 in the undergraduate dental practice, their concerns and anxieties need to be also consulted and considered. Thus, as a final remark, we suggest that our questionnaire might be a useful tool and could be applied by dental schools before returning to clinical activities.

## Data Availability

All data is available upon request

## Acknowledgment

We specially acknowledge all students who participated in this study, as well as those who engaged in our divulgation via social media. Aragao, MGB and Gomes, FIF hold FAPESP PhD. scholarships (20/02658-7 and 19/14285-3, respectively)

---

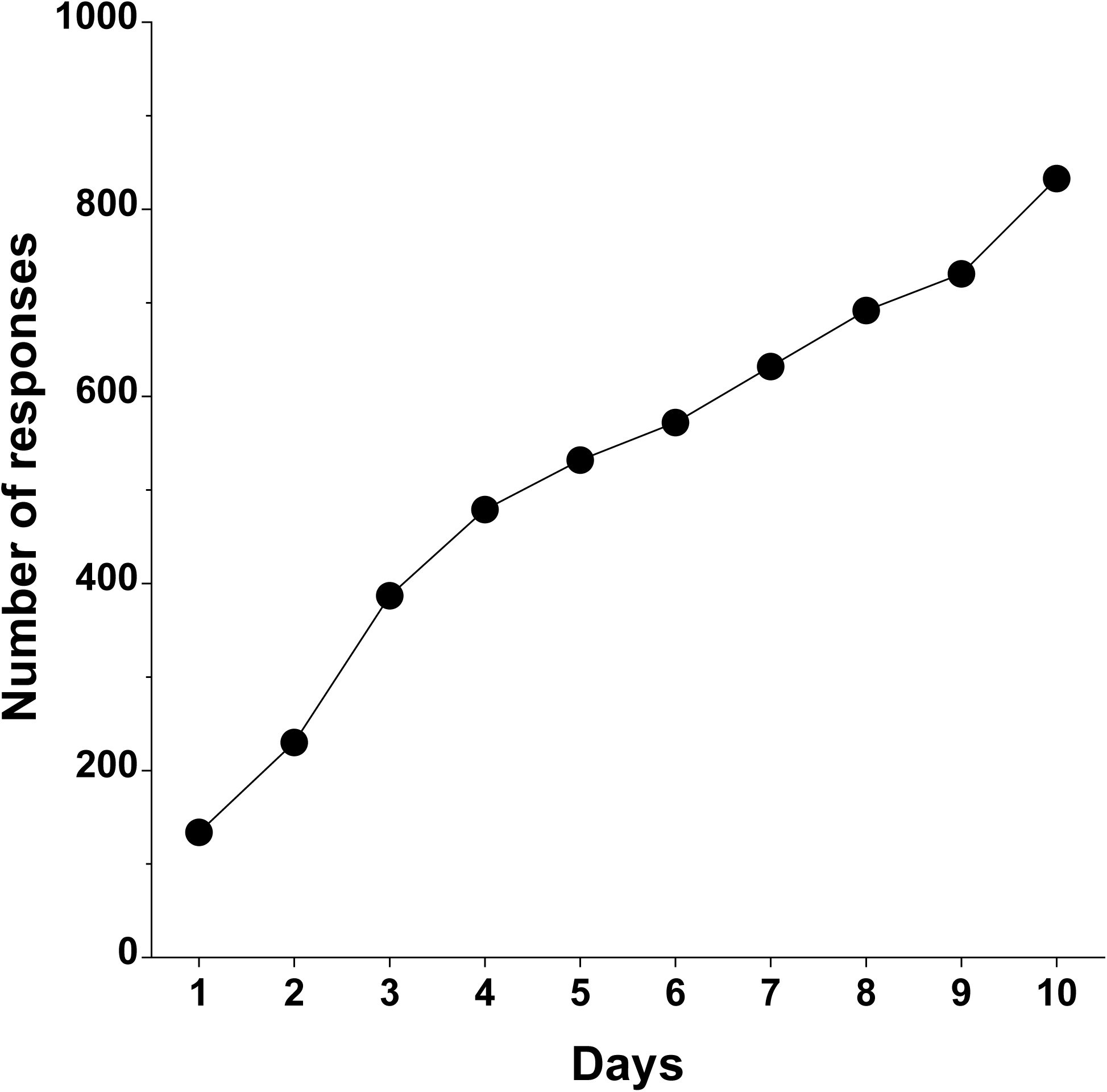

## References

Adams, J. G., & Walls, R. M. (2020). Supporting the Health Care Workforce during the COVID-19 Global Epidemic. JAMA - Journal of the American Medical Association, 323(15), 1439–1440. https://doi.org/10.1001/jama.2020.3972

Ather, A., Patel, B., Ruparel, N. B., Diogenes, A., & Hargreaves, K. M. (2020). Coronavirus Disease 19 (COVID-19): Implications for Clinical Dental Care. Journal of Endodontics, 46(5), 584–595. https://doi.org/10.1016/jjoen.2020.03.008

Baghizadeh Fini, M. (2020). What dentists need to know about COVID-19. Oral Oncology, 105(April), 104741. https://doi.org/10.1016/j.oraloncology.2020.104741

Brazilian Federal Dental Board. COVID19: Manual de Boas Práticas em BiosseguranÇa para Ambientes Odontológicos é lanÇado com apoio institucional do CFO. (n.d.). Retrieved August 5, 2020, from https://website.cfo.org.br/covid19-manual-de-boas-praticas-em-biosseguranca-para-ambientes-odontologicos-e-lancado-com-apoio-institucional-do-cfo/ (Original work published)

Brazilian National Institute of Educational Research and Studies Anisio Teixeira (INEP). Statistics of the Tertiary Education Census 2018 (n.d.). Retrieved August 5, 2020, from http://portal.inep.gov.br/web/guest/sinopses-estatisticas-da-educacao-superior

Clinical Centers for Disease Control (CDC). Guidance for Dental Settings | CDC. (n.d.). Retrieved August 5, 2020, from https://www.cdc.gov/coronavirus/2019-ncov/hcp/dental-settings.html

Couper, M. P., Traugott, M. W., & Lamias, M. J. (2001). Web survey design and administration. Public Opinion Quarterly, 65(2), 230–253. https://doi.org/10.1086/322199

Desai, B. K. (2020). Clinical implications of the COVID-19 pandemic on dental education. Journal of Dental Education, 84(5), 512. https://doi.org/10.1002/jdd.12162

Elsevier. COVID-19 Health Education resource center | Elsevier. (n.d.). Retrieved August 5, 2020, from https://www.elsevier.com/education/health-faculty-hub/covid-19

Food and Drug Administration (FDA). COVID-19 Educational Resources | FDA. (n.d.). Retrieved August 5, 2020, from https://www.fda.gov/emergency-preparedness-and-response/coronavirus-disease-2019-covid-19/covid-19-educational-resources

Ge, Z. yu, Yang, L. ming, Xia, J. jia, Fu, X. hui, & Zhang, Y. zhen. (2020). Possible aerosol transmission of COVID-19 and special precautions in dentistry. Journal of Zhejiang University: Science B, 21(5), 361–368. https://doi.org/10.1631/jzus.B2010010

Ghai, S. (2020). Are dental schools adequately preparing dental students to face outbreaks of infectious diseases such as COVID-19? Journal of Dental Education, 84(6), 631–633. https://doi.org/10.1002/jdd.12174

Gralinski, L. E., & Menachery, V. D. (2020). Return of the coronavirus: 2019-nCoV. Viruses, 12(2), 1–8. https://doi.org/10.3390/v12020135

Iyer, P., Aziz, K., & Ojcius, D. M. (2020). Impact of COVID-19 on dental education in the United States. Journal of Dental Education, 84(6), 718–722. https://doi.org/10.1002/jdd.12163

Li, Q., Guan, X., Wu, P., Wang, X., Zhou, L., Tong, Y., … Feng, Z. (2020). Early transmission dynamics in Wuhan, China, of novel coronavirus-infected pneumonia. New England Journal of Medicine, 382(13), 1199–1207. https://doi.org/10.1056/NEJMoa2001316

Liu, X., Zhou, J., Chen, L., Yang, Y., & Tan, J. (2020). Impact of COVID_ 19 epidemic on live online dental continuing education. European Journal of Dental Education, (June), 1–4. https://doi.org/10.1111/eje.12569

Machado, R. A., Bonan, P. R. F., Perez, D. E. da C., & Martelli JUnior, H. (2020). COVID-19 pandemic and the impact on dental education: discussing current and future perspectives. Brazilian Oral Research, 34, e083. https://doi.org/10.1590/1807-3107bor-2020.vol34.0083

Meng, L., Hua, F., & Bian, Z. (2020). Coronavirus Disease 2019 (COVID-19): Emerging and Future Challenges for Dental and Oral Medicine. Journal of Dental Research, 99(5), 481–487. https://doi.org/10.1177/0022034520914246 (Original work published)

Moraes, R. R., Correa, M. B., Queiroz, A. B., Daneris, A., Lopes, J. P., Pereira-Cenci, T., … Demarco, F. F. (2020). COVID-19 challenges to dentistry in the new pandemic epicenter: Brazil. MedRxiv, 2020.06.11.20128744. https://doi.org/10.1101/2020.06.11.20128744

Omolo, C. A., Soni, N., Fasiku, V. O., Mackraj, I., & Govender, T. (2020). Update on therapeutic approaches and emerging therapies for SARS-CoV-2 virus. European Journal of Pharmacology, 883(June), 173348. https://doi.org/10.1016/j.ejphar.2020.173348

Peng, X., Xu, X., Li, Y., Cheng, L., Zhou, X., & Ren, B. (2020). Transmission routes of 2019-nCoV and controls in dental practice. International Journal of Oral Science, 12(1), 1–6. https://doi.org/10.1038/s41368-020-0075-9

Prati, C., Pelliccioni, G. A., Sambri, V., Chersoni, S., & Gandolfi, M. G. (2020). COVID-19: its impact on dental schools in Italy, clinical problems in endodontic therapy and general considerations. International Endodontic Journal, 53(5), 723–725. https://doi.org/10.1111/iej.13291

Qian, M., & Jiang, J. (2020). COVID-19 and social distancing. Journal of Public Health (Germany), (Mikulska 2019). https://doi.org/10.1007/s10389-020-01321-z

Quinn, B., Field, J., Gorter, R., Akota, I., Manzanares, M. C., Paganelli, C., … Tubert-Jeannin, S. (2020). COVID-19: The Immediate Response of European Academic Dental Institutions and Future Implications for Dental Education. European Journal of Dental Education, (May), 1–4. https://doi.org/10.1111/eje.12542

Raimondi, M. T., Donnaloja, F., Barzaghini, B., Bocconi, A., Conci, C., Parodi, V., … Carelli, S. (2020). Bioengineering tools to speed up the discovery and preclinical testing of vaccines for SARS-CoV-2 and therapeutic agents for COVID-19. Theranostics, 10(16), 7034–7052. https://doi.org/10.7150/thno.47406

Sahu, P. (2020). Closure of Universities Due to Coronavirus Disease 2019 (COVID-19): Impact on Education and Mental Health of Students and Academic Staff. Cureus, 2019(4), 4–9. https://doi.org/10.7759/cureus.7541

UNA-SUS. Especial Coronavirus (COVID-19) - UNA-SUS. (n.d.). Retrieved August 5, 2020, from https://www.unasus.gov.br/especial/covid19/

United Nations Educational Scientific Cultural Organization (UNESCO). School closures caused by Coronavirus (Covid-19). (n.d.). Retrieved August 5, 2020, from https://en.unesco.org/covid19/educationresponse (Original work published)

Van Doren, E. J., Lee, J. E., Breitman, L. S., Chutinan, S., & Ohyama, H. (2020). Students’ perceptions on dental education in the wake of the COVID-19 pandemic. Journal of Dental Education, (June), 3–5. https://doi.org/10.1002/jdd.12300

Wang, C., Horby, P. W., Hayden, F. G., & Gao, G. F. (2020). A novel coronavirus outbreak of global health concern. The Lancet, 395(10223), 470–473. https://doi.org/10.1016/S0140-6736(20)30185-9

We Are Social. Digital use around the world in July 2020 (n.d.). Retrieved August 5, 2020, from https://wearesocial.com/blog/2020/07/digital-use-around-the-world-in-july-2020

WHO Coronavirus Disease (COVID-19) Dashboard | WHO Coronavirus Disease (COVID-19) Dashboard. (n.d.). Retrieved August 5, 2020, from https://covid19.who.int/

World Health Organization (WHO). Clinical management of COVID-19. (n.d.). Retrieved August 5, 2020, from https://www.who.int/publications/i7item/clinical-management-of-covid-19

World Health Organization (WHO). Clinical management of COVID-19: interim guidance (n.d.). Retrieved August 8, 2020, from https://apps.who.int/iris/handle/10665/332196

Wu, D. T., Wu, K. Y., Nguyen, T. T., & Tran, S. D. (2020). The impact of COVID-19 on dental education in North America—Where do we go next? European Journal of Dental Education, (March), 1’3. https://doi.org/10.1111/eje.12561

